# Health and economic impact of the human papillomavirus (HPV) vaccine in Mozambique

**DOI:** 10.1101/2025.06.17.25329752

**Authors:** Paula Christen, Agostinho Viana Lima, Muanacha Mintade, Neusa Torres, Timothy B. Hallett, Lesong Conteh, Allison Portnoy

## Abstract

2

Mozambique is among the countries with the highest cervical cancer (CC) incidence rates in the world. Yet, in sub-Saharan Africa, only 5% of CC patients receive appropriate treatment. In 2021, Mozambique integrated the prophylactic human papillomavirus (HPV) vaccine into its national immunization program as a primary prevention strategy. As part of a qualitative research effort on participatory modeling, this study aimed to quantify the magnitude and distribution of expected health and economic outcomes of the current nationwide HPV vaccination program, hypothetical scale-up scenarios, and global targets.

We applied and extended a closed cohort model to quantify the vaccine program’s direct potential population health benefits (in averted CC cases and deaths), financial savings to patients (in direct medical treatment costs saved and in financial risk protection provided), health provider benefits measures in saved labour (full-time equivalents, FTEs) and distribution of benefits across the entire country under empirically informed and potential vaccination coverage scenarios.

With a coverage of 18.9% in 2021, the vaccination program is estimated to avert up to 9.7% of expected CC cases and deaths (from 4.0% in the highest socioeconomic quintile (HQ) to 11.6% in the lowest socioeconomic quintile (LQ)). Across the lifetime of a single vaccination cohort, an average of US$106,532 (min: US$48,382, max: US$157,811) in direct medical CC treatment expenditure for patients and the time equivalent to 3.3 (2.6 – 3.9) full-time healthcare workers is saved per year. Over 30 years, 50% vaccination coverage could prevent 23.9% of predicted cases and fatalities (LQ: 25.2% to HQ: 14.8%). With a gradient in vaccine coverage favoring higher socioeconomic quintiles in urban areas, the benefits in the LQ rural population are estimated to be six times greater than in the LQ urban population.

These findings can contribute to fair and efficient planning decisions from various stakeholder perspectives in policy dialogues around the HPV vaccination programme in Mozambique, and at a global level.

**Key messages:**

- This study evaluates the health and economic effects of Mozambique’s HPV vaccination program using a cohort-based modeling approach, estimating reductions in cervical cancer cases, financial burden, and healthcare system demands.
- At 50% vaccine coverage, the program could avert 26% of expected cervical cancer cases and deaths, with the highest impact observed among lower-income populations who face greater disease burden and limited access to care.
- The financial benefits extend beyond individual health gains, as the program could reduce catastrophic health expenditures, alleviate pressure on the healthcare workforce, and contribute to more equitable health outcomes.
- These insights support strategic decision-making on HPV vaccination expansion, highlighting the need for targeted efforts to maximize both effectiveness and equity in Mozambique’s cancer prevention initiatives.

## 4 Introduction

Cervical cancer (CC), primarily caused by persistent infection with high-risk human papillomavirus (HPV) types, remains a significant health burden globally, disproportionately affecting women in low- and middle-income countries (LMICs) (De Martel *et al*., 2017). In Mozambique, CC is the leading cause of cancer death (International agency for research on cancer - The Global Cancer Observatory, 2022). In 2020, Mozambique had one of the world’s highest estimated age-standardized CC incidence rates (50.2 per 100,000), 3.7 times higher than the global average (13.3 per 100,000) and 2.4 times higher than the southern African regional estimate (20.6 per 100,000) (Sung *et al*., 2021). Like many LMICs, Mozambique faces significant challenges in CC prevention and control due to limited healthcare resources (Lorenzoni *et al*., 2018; Tulsidás *et al*., 2023), low screening coverage (Brandão *et al*., 2019), and high treatment costs (Omotoso *et al*., 2023).

The introduction of the HPV vaccine in November 2021 presents a promising strategy to reduce the health and economic burden of CC in Mozambique. However, the success of the vaccination programme depends on vaccine coverage, health system capacity, and socioeconomic factors. Disparities in disease burden and vaccine uptake across socioeconomic groups can significantly shape its impact on health equity (Sacre *et al*., 2023; Abbas *et al*., 2024). Despite efforts to ensure equitable healthcare access, marginalized groups remain underserved (Ali *et al*., 2022). Socioeconomic and behavioral risk factors - including lower education (Singini *et al*., 2021; Onwuamah *et al*., 2023) and income levels (Berraho *et al*., 2017; Abila *et al*., 2021) - exacerbate CC risk and HPV infection rates. Within-country disparities in education, healthcare access, and economic stability have also been shown to affect HPV vaccine uptake in Mozambique (Haidari *et al*., 2017). Mozambique rolled out HPV vaccination nationwide primarily via school-based campaigns, complemented by door-to-door and mobile outreach to reach out-of-school adolescents (World Health Organization, 2024).

Reaching underserved populations remains a challenge, particularly for slum residents and nomadic groups in the East African region (Watson-Jones *et al*., 2015; Bonner, Banura and Basta, 2018; Kutz *et al*., 2023). School-based delivery systems, while effective, may inadvertently exclude out-of-school adolescents and those in private schools (Lubeya *et al*., 2024). Furthermore, focusing solely on adolescents overlooks other high-risk groups who could benefit from vaccination (Piñeros *et al*., 2010). Similar urban-rural disparities in CC incidence further underscore the need for equity-focused vaccination strategies (Berraho *et al*., 2017; Singini *et al*., 2021).

Beyond health benefits, HPV vaccination offers financial protection. CC treatment is costly, with screening and treatment in Tanzania averaging US$2,526 (Nelson *et al*., 2016). This substantial financial burden, which is not covered by the government (Das, 2022), leads to high out-of-pocket expenditures, increasing the risk of financial hardship. In contrast, the HPV vaccine is a relatively low-cost intervention (approximately US$6 per dose delivered in a Mozambican pilot programme (Alonso *et al*., 2019)) and is largely funded by international support – for instance, about 80% of Mozambique’s immunization programme costs are covered by Gavi, the Vaccine Alliance (Gavi, the Vaccine Alliance, 2025).

In addition to reducing CC cases, the HPV vaccination programme could alleviate pressure on the healthcare system (Guillaume *et al*., 2023). Many African countries face a shortage of healthcare workers, particularly cancer surgeons (Sheffel *et al*., 2024). Preventing CC cases reduces the demand for cancer treatment, helping mitigate workforce shortages (Brassel *et al*., 2023). With CC incidence projected to rise in LMICs over the next 50 years - driven by population growth, aging, urbanization, and lifestyle changes (Soerjomataram and Bray, 2021) - ensuring sustainable healthcare workforce capacity is crucial. In Mozambique, healthcare workforce costs constitute 42% of non-communicable disease interventions (Belardi, Nollino and Putoto, 2024).

The objective of this study was to assess the impact of current HPV vaccination programme efforts, hypothetical scale-up scenarios, and global targets on population health, healthcare workforce, financial risk, and health disparities in Mozambique. We evaluated the magnitude and distribution of expected health and economic outcomes across scenarios that assume increased coverage of the HPV vaccine and/or government-funded treatment coverage.

## 5 Methods

We used the Extended Cost-Effectiveness Analysis (ECEA) framework to evaluate the impact of the current HPV vaccination program, targeting 9-year old girls, hypothetical scale-up scenarios, and the global coverage target (90%) on population health, the healthcare workforce, financial risk, and health disparities (Verguet, Kim and Jamison, 2016). We evaluated the vaccination programme from a health sector perspective.

### 5.1 The model and its biological assumptions

To estimate health gains, we applied a “closed” static cohort model (Goldie *et al*., 2008), an epidemiological modeling approach that tracks a defined population over time without accounting for transmission to or from the cohort. This model estimates aggregate health outcomes, which were then stratified by population subgroups. Following the ECEA approach, the model enables the quantification of health benefits across subgroups by accounting for variations in (i) vaccine coverage across different population groups, and (ii) disease burden distribution among these groups. The estimated health gains from the cohort model served as input values to assess benefits from a health system perspective, including the distribution of health improvements and financial risk protection across population subgroups. Figure 1 provides an overview of how the model is applied within the ECEA framework. Assumptions and data sources are presented in Table 1.

**Figure 1.**
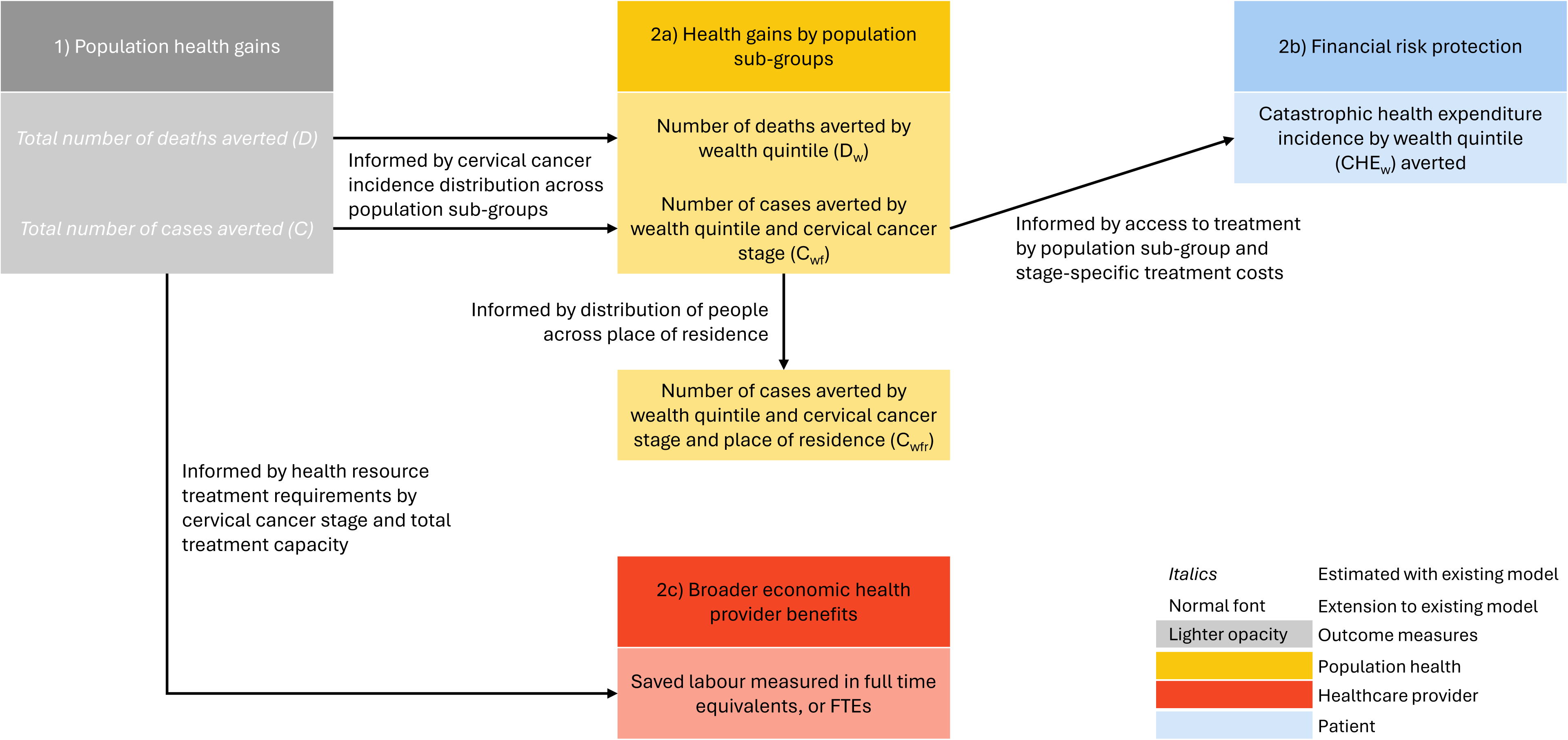
Schematic of the extended cost effectiveness analysis (ECEA) approach application. Population health gains (grey box) are estimated using Goldie et al.’s static cohort model. All other outcomes are computed with outcomes from Goldie et al.’s model.

**Table 1.** Parameter values (at baseline).

| Parameter | Value | Reference |
| --- | --- | --- |
| **Female population in Mozambique** | Total female population as of 2022: 16,793,302 | (United Nations Department of Economic and Social Affairs Population Division, 2022) |
| **CC incidence by age** | See Table 1 in supplement for cervical cancer incidence per 100,000 | (Union for International Cancer Control, 2019) |
| **HPV-16/18 type distribution** | 0.51/0.49 | (Edna Omar *et al.*, 2017) |
| **Stage distribution of detected cancers by FIGO stage** | Stage I: 0.09<br>Stage II: 0.36<br>Stage III: 0.47<br>Stage IV: 0.08 | (Carrasco-Escobar, Fornace and Benmarhnia, 2021) |
| **Vaccination coverage** | 18.9% | (Anonymous Interviewee, 2023) |
| **Vaccine wastage** | 5% | (Parmar *et al.*, 2010) |
| **One-time-introduction cost per targeted girl in US\$** | 2.00 | (Portnoy *et al.*, 2021) |
| **Recurrent delivery cost per girl in US\$** | 1.70 | (Portnoy *et al.*, 2021) |
| **Vaccine price per dose in US\$** | 4.50 | (Portnoy *et al.*, 2021) |
| **Distribution of CC incidence across wealth quintiles** | Q1: 29.7%<br>Q2: 20.9%<br>Q3: 24.7%<br>Q4: 13.9% | (Rweyemamu, 2013) |
|  | Q5: 10.8% |  |
| **Distribution of wealth quintiles across residence types** | Urban Q1: 57%<br>Urban Q2: 27%<br>Urban Q3: 9%<br>Urban Q4: 3%<br>Urban Q5: 4%<br>Rural Q1: 27%<br>Rural Q2: 28%<br>Rural Q3: 25%<br>Rural Q4: 17%<br>Rural Q5: 3% | (Instituto Nacional de Estatística, 2021) |
| **Distribution of population across types of residence** | Urban: 38%<br>Rural: 62% | (Instituto Nacional de Saúde and ICF/Programa DHS, 2015) |
| **Direct medical treatment costs by FIGO stage in US\$** | Stage I: 2,989<br>Stage II: 2,999<br>Stage III: 1,744<br>Stage IV: 1,741 | (Nelson *et al.*, 2016) |
| **Proportion of direct medical treatment costs covered by healthcare provider** | 0 | (Das, 2022) |
| **CHE threshold** | 10% of annual household expenditure | (Eze *et al.*, 2022) |
| **Average household size** | 4.78 | (Instituto Nacional de Estatística, 2021) |
| **Monthly per capita consumption in MZN (2019/20)** | Q1: 224<br>Q2: 585<br>Q3: 990<br>Q4: 1,655<br>Q5: 5,022 | (Instituto Nacional de Estatística, 2021) |
| **Proportion of patients seeking treatment at Central Private Hospital** | 4.70% | (Gironés *et al.*, 2020) |
| **Access to general care by wealth quintile** | Q1: 57.0%<br>Q2: 67.8%<br>Q3: 66.6%<br>Q4: 79.9%<br>Q5: 81.8% | (Instituto Nacional de Estatística, 2021) |
| **Medical personnel labour cost by FIGO stage in US\$** | Stage I: 106.95<br>Stage II: 117.39<br>Stage III: 105.16<br>Stage IV: 102.24 | (Nelson *et al.*, 2016) |
| **Annual treatment capacity for CC patients at public hospital** | 900 | (‘Prevention, diagnosis and management of cancer in Mozambique’, 2021) |
| **Demand for CC treatment (2021)** | 5,325 | (Bray *et al.*, 2018) |

### 5.2 Health gain distribution

To stratify health outcomes by socioeconomic subgroup and residence (urban and rural), we assumed that the distribution of cervical cancer incidence across wealth quintiles in Mozambique mirrors that observed in Tanzania (Umuhoza and Ataguba, 2018), given the absence of Mozambican data. Tanzania provides a reasonable proxy as it has a comparable disease burden (age-standardised incidence rate of 62.5 vs. 50.2 per 100,000 women) (Bruni *et al*., 2023) and nationally representative evidence of steep pro-rich, pro-urban screening gradients (Abila *et al*., 2024; Asgedom *et al*., 2024; Mahiti, Adam and Luoga, 2025). While Mozambique’s higher HIV prevalence, deeper poverty, and greater inequality likely mean the true gradient is even steeper, this assumption offers a pragmatic baseline for analysis.

To do this for residence type, it was used data from Mozambique on the distribution of persons in each wealth quintile by urban and rural residence (Instituto Nacional de Estatística, 2021).

### 5.3 Additional model outcomes

The model was also used to estimate **vaccination programme costs**. We accounted for vaccine wastage, one-time introduction, and recurrent delivery costs (Table 1), following Portnoy, Sweet, et al., 2021. The one-time introduction cost estimate includes non-recurrent introduction activities, i.e., planning, training, social mobilization, and information, education, communication in the first year, while the recurrent cost estimate includes ongoing costs for service delivery. All future costs were discounted annually at a rate of 3%.

**Health resource requirements** were derived from population health outcomes using a needs-based workforce modelling approach (Asamani, Christmals and Reitsma, 2021). With this approach, the number of health professionals, quantified in full-time equivalents (FTE) required to meet a population’s health needs was estimated based on the estimated prevalence of the disease and the appropriate standards of care for patients with the disease.

**Healthcare need** is defined by the number of CC patients by cancer stage per year. Capacity is a function of labour productivity. Labour and productivity estimates from Nelson et al.’s (Nelson *et al*., 2016) study were used to derive the number of hours required to treat CC patients by cancer stage. In Mozambique, full time medical staff works 210 days per year, 8 hours a day (Dutta *et al*., 2014).

Health outcomes by population subgroup are used to estimate catastrophic health expenditure (CHE) incidence. CHE, defined as out-of-pocket spending that exceeds a given income or consumption threshold, is a key metric for assessing financial risk (Eze *et al*., 2022). For each wealth quintile, CHE incidence was determined by the number of CC cases by stage and wealth quintile, access to CC treatment by wealth quintile, stage-specific direct treatment costs, the proportion of direct medical expenditure financed out of pocket, and annual household expenditure. Supplement 1 provides more information on how parameter values were derived.

### 5.4 Model scenarios

To evaluate the potential impact of the HPV vaccination program, we modeled several scenarios reflecting different vaccination coverage levels and distribution patterns. The analysis includes a base-case scenario (A) that relies on current vaccination coverage and treatment cost assumptions, scale-up scenarios (C and D) that examine uniform increases in coverage, and coverage distribution scenarios (D and E) that account for variations by wealth quintile and urban versus rural residence. Assumptions for each scenario are provided in Table 2.

**Table 2.** Analytical scenarios modelled.

| Scenario label | Scenario | Description | Coverage | Source for assumption |
| --- | --- | --- | --- | --- |
| **A** | Base case | Informed by vaccination coverage, assumed to be uniformly distributed across the population, recorded in the first year of the programme (2021). | 18.9% | Anonymous Interviewee, 2023 |
| **B** | Global | Informed by global Cervical | 90% | Cervical Cancer |
|  | target | Cancer Elimination Initiative target. |  | Elimination Initiative's goal |
| **C** | Half-way, flat coverage | Vaccination coverage is uniformly distributed across wealth quintiles. | 50% |  |
| **D** | Half-way, gradient coverage by quintile | The vaccination programme is assumed to have a gradient coverage across wealth quintiles, with an overall weighted vaccine coverage average of 50% across the population. The gradient is empirically informed by access to general healthcare. | Q1: 41.2%<br>Q2: 40.6%<br>Q3: 44.7%<br>Q4: 55.3%<br>Q5: 68.2% | Instituto Nacional de Estatística, 2021 |
| **E** | Half-way, gradient coverage by residence | The vaccination programme is assumed to have a gradient coverage across populations living in different areas of residence (Instituto Nacional de Saúde and ICF/Programa DHS, 2015), with an overall weighted vaccine coverage average of 50% across the population. The gradient is empirically informed by access to general healthcare. | Urban: 63.9%<br>Rural: 36.1% | Instituto Nacional de Estatística, 2021 |

### 5.5 Analysis plan

Outcome measures and sensitivity analysis are summarized in Table 3.

**Table 3.**
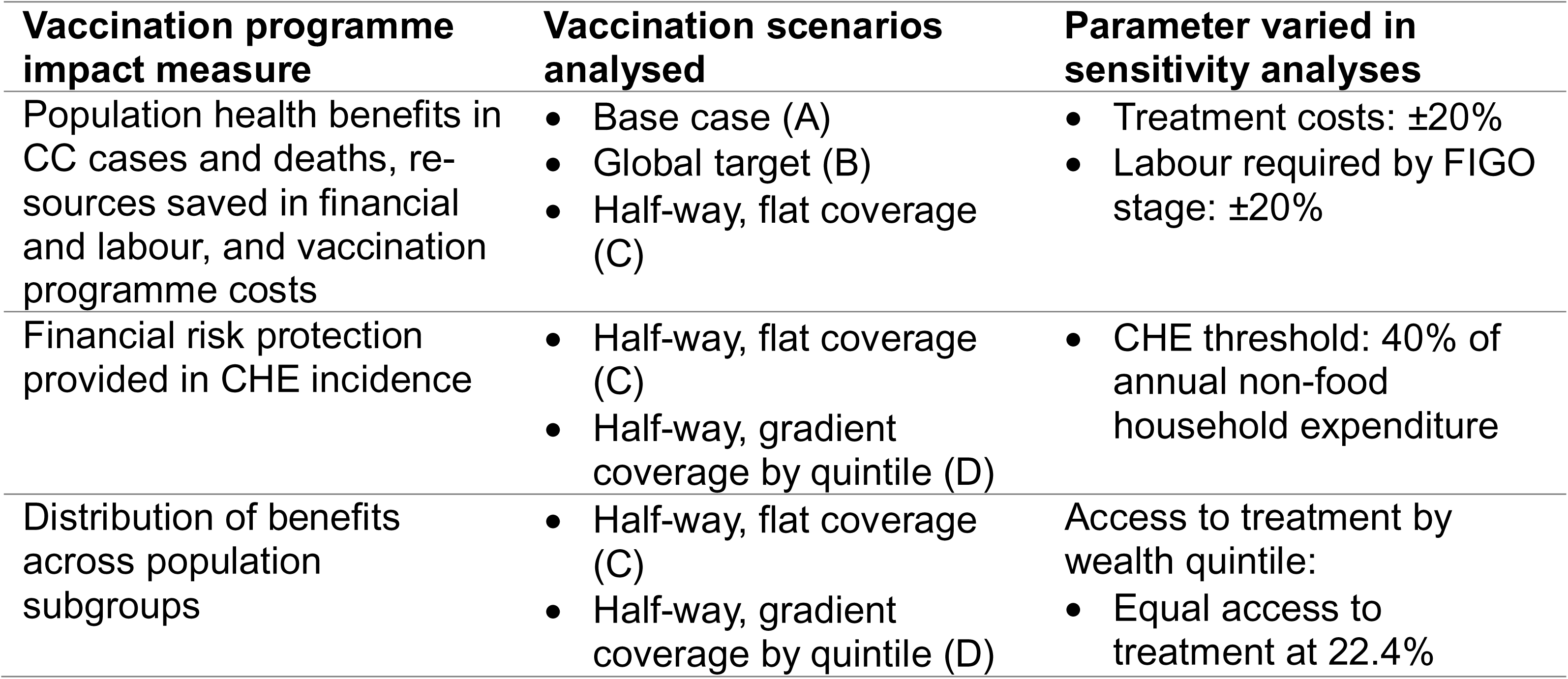
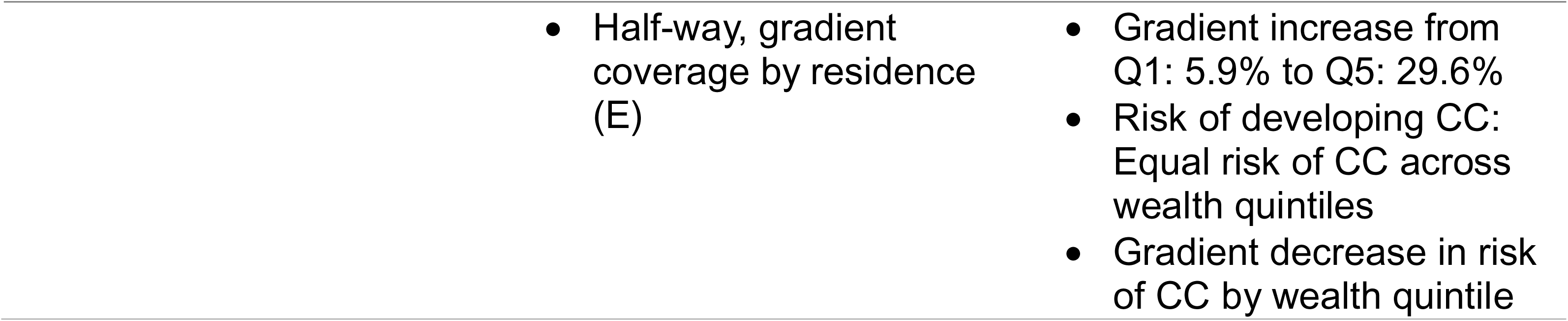
Outcome measures by analytic scenario.

Population health benefits and resources (labour) saved attributable to the vaccination programme were quantified by estimating the number of CC cases and deaths, as well as vaccination programme costs saved. Further, these outcomes were estimated under the base case scenario (A) and compared to health benefits estimated with the hypothetical scale-up scenarios (B, C). Health benefits estimated in scenario C were compared with health benefits estimated in scenario D.

The difference in CHE incidence between scenarios A and C was quantified and CHE incidence reduction attributable to the vaccination programme was computed. CHE was defined as the number of out-of-pocket spending incidences exceeding 10% of total consumption or income (Eze *et al*., 2022).

We also compared outcomes of scenarios A and D. We examined how the effect is influenced by alternative assumptions about the access to treatment afforded to people in different income quintiles.

We also explored what proportion of treatment costs would have to be covered by a public finance scheme to reduce CHE incidence, in addition to the vaccination programme.

One-way sensitivity analyses were conducted by varying five key parameters over plausible ranges. Treatment costs and labour requirements in terms of FTE were adjusted by ±20% for each cervical cancer (CC) stage. To assess the impact of different financial risk protection definitions, an alternative CHE threshold was explored by setting it at 40% of non-food household expenditure. Non-household food expenditure was informed by the Budget Household Survey 2019/2020 (Instituto Nacional de Estatística, 2021). Access to treatment was examined under two assumptions: equal access across all wealth quintiles and a gradient increase in access from the poorest to the richest quintile (ranging from 5.9% in the poorest group to 29.6% in the wealthiest, with an average of 22.4%), based on observed trends in Nigeria and Uganda (Foerster *et al*., 2019). Finally, CC incidence distribution was tested under two scenarios: one assuming an equal probability of CC across wealth quintiles, and another assuming a lower probability of CC in higher wealth quintiles, modifying the initial distribution based on registry data from Dar es Salaam, Tanzania (Rweyemamu, 2013).

We note that several input parameters relied on proxy or partial data due to gaps in national statistics. For example, age-specific incidence rates are based on registries from only two hospitals (Beira and Maputo), and the distribution of cervical cancer incidence across wealth quintiles was adapted from Tanzanian data in the absence of Mozambican evidence. Such constraints mean that while the model provides robust directional insights, absolute estimates should be interpreted with caution. Full details are explored in the Discussion.

## 6 Results

### 6.1 Health impact

Across the lifetime of a single vaccinated cohort, approximately 1,700 cervical cancer cases and 600 cervical cancer deaths (10%) are estimated to be averted. The scale of impact varies by coverage scenario. Under Scenario C, up to 26% of cervical cancer cases and deaths would be prevented, while under Scenario B, the reduction would be 46%.

### 6.2 Health system impact

Total vaccination programme costs were estimated at US$1,080,184, which represents 49% (95% uncertainty interval: 39% – 59%) of the medical treatment costs saved by vaccinating 18.9% of females in the target group. The vaccination programme under Scenario B is associated with costs of US$113M, with cost savings proportionate to vaccination coverage and the medical treatment costs averted.

The estimated labor saved due to vaccination was equivalent to the mean annual full-time workload of 3.3 (2.6 – 3.9) FTEs under Scenario A. Under Scenario B, an estimated 780,000 hours (624,000 – 936,000) of medical labor would be saved; for one year this corresponds to 15.5 (12.4 – 18.6) FTEs. Under Scenario D, the labor saved would amount to 8.0 (6.4 – 9.6) FTEs per year.

### 6.3 Household budget impact

The vaccination programme is expected to significantly reduce CHE related to cervical cancer treatment. With 100% of direct medical costs financed out of pocket, approximately 6,000 cases of CHE and more than US$100M in treatment costs could be averted over the lifetime of girls across 30 cohorts, i.e., every target cohort of 9-year-old girls from 2021 to 2051. If vaccination coverage is higher among wealthier population sub-groups, as likely to be the case, the number of CHE cases averted would be reduced by 200 persons.

If CHE is measured using a threshold of 40% of non-food annual household expenditure, the incidence would be similar to estimates using a 10% threshold of total annual household expenditure. If the CHE threshold is defined as 10% of annual household expenditure and a public financing scheme in Mozambique were to cover 76% of direct medical treatment costs, the incidence of CHE would be reduced by 10%. Ninety-six percent of direct medical treatment costs would need to be covered to avert 51% of CHE incidence. If CHE is defined using a threshold of 40% of non-food annual household expenditure, public financing covering 26% of direct medical treatment costs would reduce CHE incidence by 11%, whereas 92% coverage of direct medical costs would be required to avert 51% of CHE cases.

**Figure 2.**
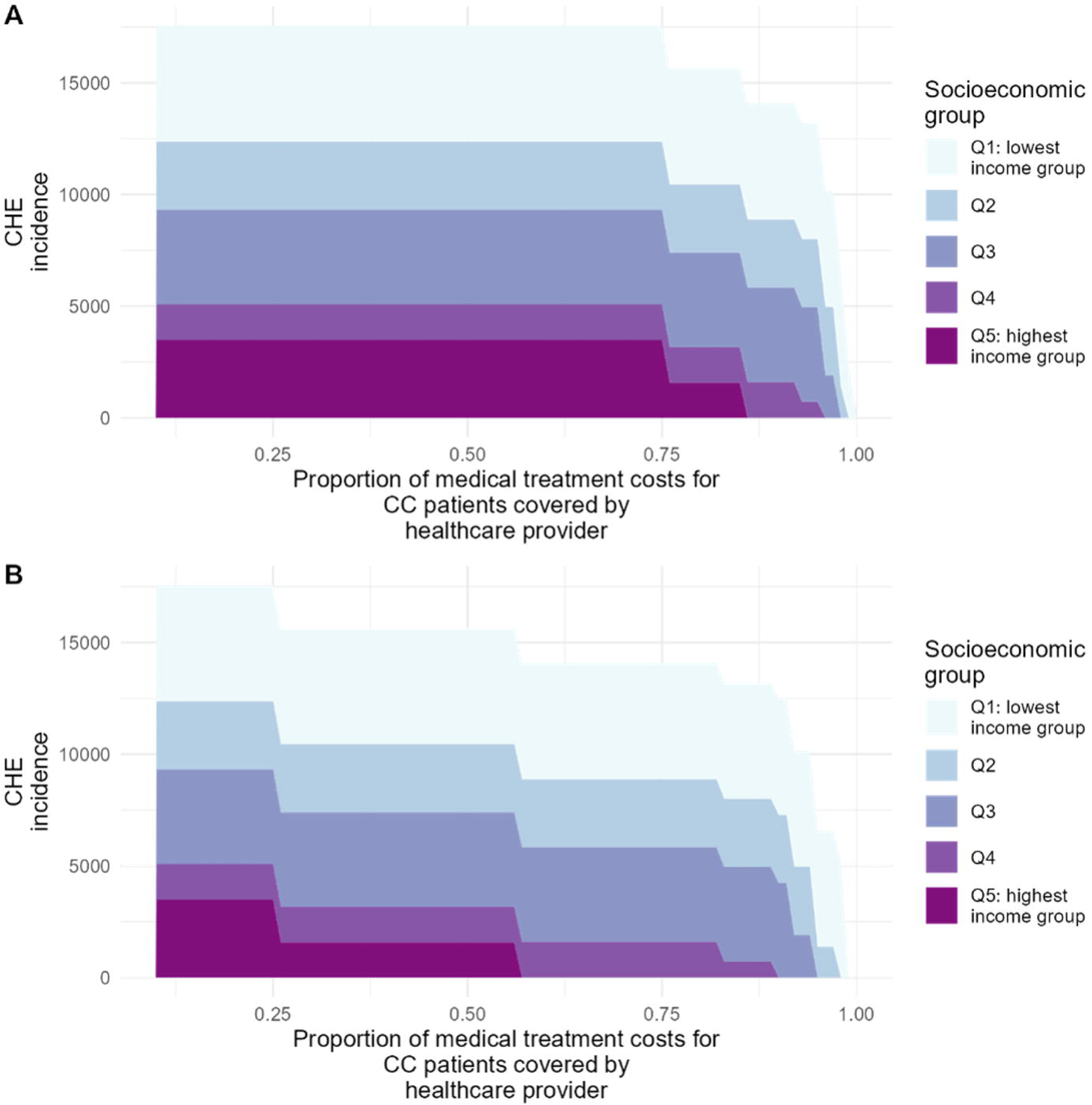
Catastrophic health expenditure (CHE) incidence with increasing medical treatment coverage for cervical cancer patients by socio-economic quintile with a threshold of (A) 10% of annual household consumption, and (B) 40% of annual non-food household consumption.

### 6.4 Distributional impact

The distribution of benefits from the vaccination programme varies across population sub-groups. Under Scenario D, the primary beneficiaries are individuals in the two lowest-income quintiles, with the poorest quintile receiving 26% of the total benefits, followed closely by the second-poorest quintile at 23%. Overall, 68% of benefits accrue to the lowest, lower-middle, and middle-income quintiles, representing the bottom 60% of the population. If the risk of cervical cancer were equal across all wealth quintiles, the highest number of cases averted would occur in the wealthiest group due to higher vaccination coverage.

**Figure 3.**
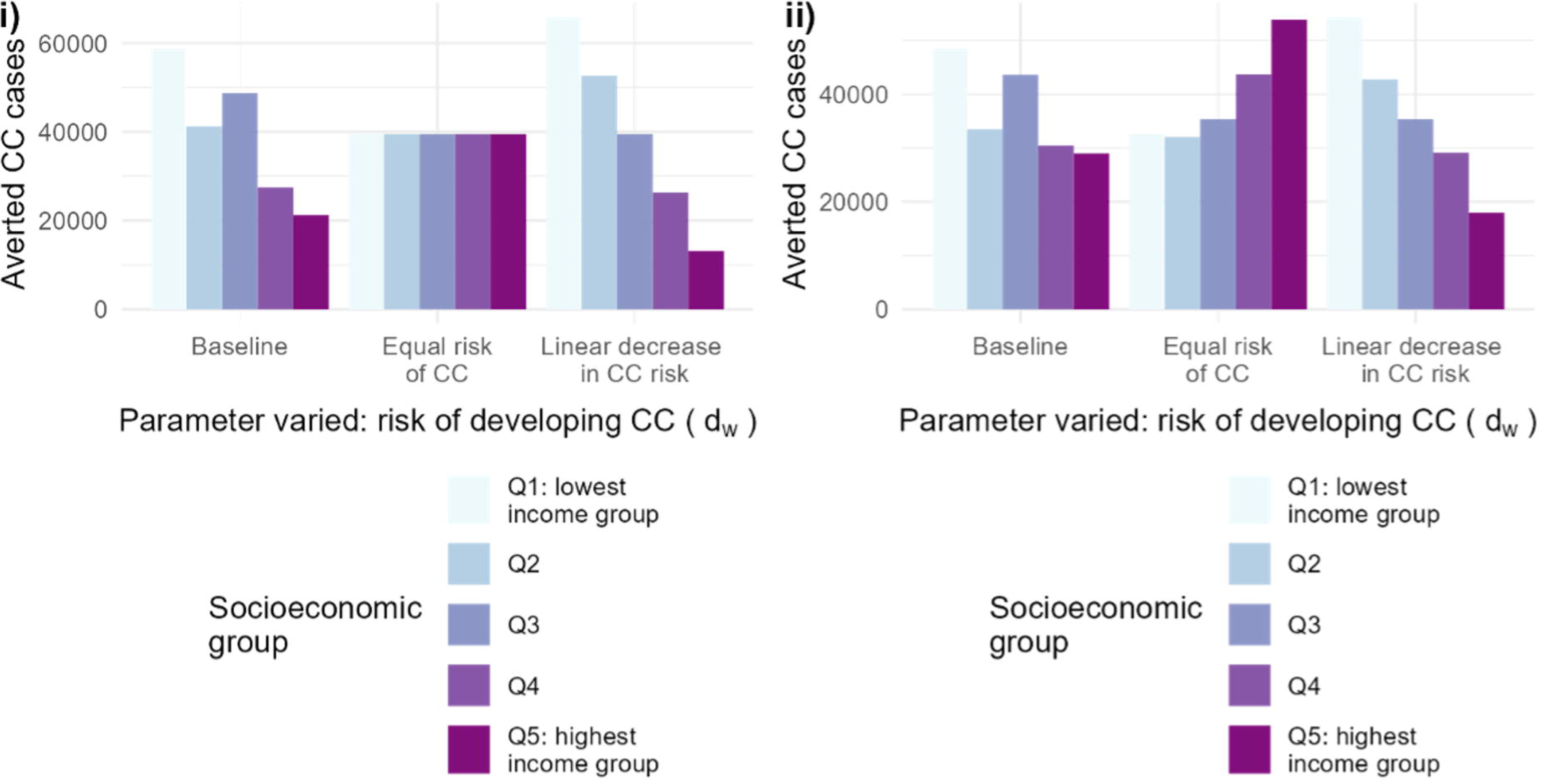
Predicted number of averted cases by socio-economic quintile under different cervical cancer (CC) risk distribution across wealth quintiles by vaccination coverage scenario. (i) 50% coverage of the HPV vaccine across wealth quintiles, and (ii) 50% of coverage of the HPV vaccine across wealth quintiles, and (ii) 50% of coverage at an increasing gradient from lowest to highest socio-economic quintile.

Under Scenario C, 30% of expected benefits would be concentrated in the lowest income quintile, and the highest number of CHE cases would be averted in this group. Estimated health gains under Scenario E would primarily benefit the highest-income urban population due to greater access to treatment, where 7% of expected cervical cancer cases would be prevented. In rural areas, the largest health benefits would accrue to the second-lowest income quintile, with an estimated 3% of expected cervical cancer cases averted.

As expected, if individuals in the poorest socio-economic quintile have limited access to treatment, the highest number of CHE cases averted would be among the highest-income groups under both Scenarios C and D, regardless of the CHE threshold definition applied.

**Figure 4.**
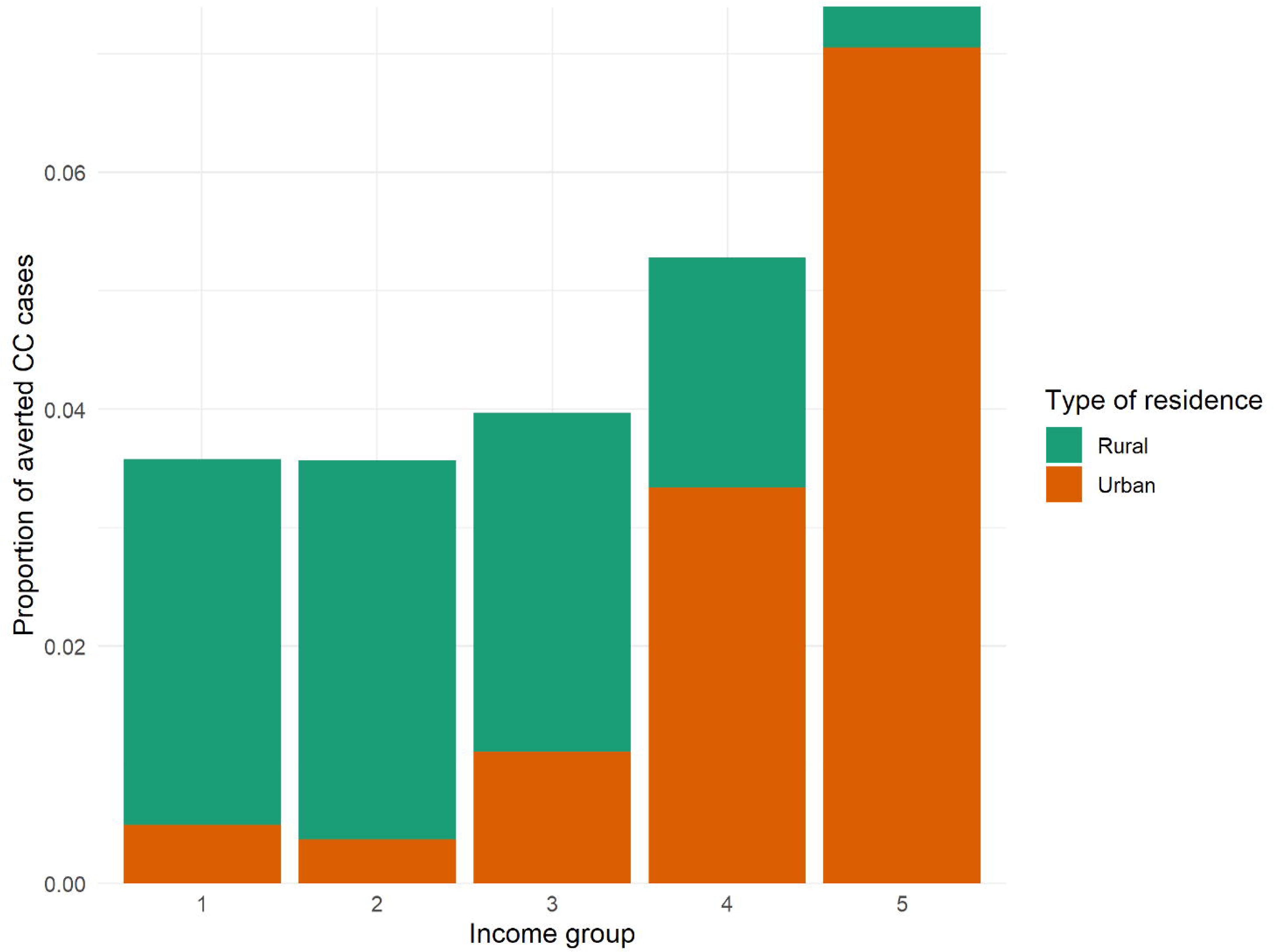
Estimated averted cervical cancer (CC) cases in percentage by wealth quintile and type of residence assuming a vaccination coverage gradient across population sub-groups living in different types of residence.

## 7 Discussion

This study applied an established model of HPV, cervical cancer (CC), and the Extended Cost-Effectiveness Analysis (ECEA) framework to evaluate the impact of Mozambique’s recently introduced HPV vaccination program from both a patient and health system perspective. The findings highlight that while the vaccination program has the potential to substantially improve population health outcomes and provide financial risk protection, its benefits are not distributed equally across socioeconomic groups.

The concentration of benefits among wealthier populations is primarily driven by the correlation between lower income, greater baseline risk of CC, and limited access to care. Even when vaccine distribution is equal, disparities in disease burden and healthcare access mean that wealthier populations experience greater relative gains. Ensuring equitable benefits would require targeted efforts to improve uptake in rural and lower-income populations. This reflects patterns observed in other settings. In sub-Saharan Africa, lower-income women generally face a higher CC burden but have less access to preventive services, creating an inequity in outcomes (Seifu *et al*., 2025). Even if vaccine distribution is equal, socio-economic disparities can lead to unequal gains. Wealthier or urban populations tend to have better healthcare access and awareness, which translates into higher uptake and more effective use of new interventions (Kutz *et al*., 2023; Agimas *et al*., 2024). For example, a meta-analysis in East Africa found girls from higher-wealth households were significantly more likely to receive the HPV vaccine than those from the poorest families (Agimas *et al*., 2024). This aligns with the broader “inverse equity” phenomenon in global health, where initial phases of new interventions often disproportionately benefit the better-off until coverage expands to disadvantaged groups (Lee *et al*., 2015). As a result, without deliberate efforts to reach low-income and rural communities, the early gains of vaccination programs can skew pro-rich, even though poorer women bear a greater disease burden.

The analysis also suggests that reducing out-of-pocket payments for CC treatment could mitigate CHE, but significant financial protection would only be achieved if treatment costs were predominantly subsidized by the state – particularly for individuals in the lowest socioeconomic quintile. In practice, only fully state-funded treatment would reduce CHE incidence by about half, and the true financial burden is likely even greater, since non-medical costs borne by patients and families (e.g., transport and food) were not captured in this analysis. To put this in perspective, meta-analysis across sub-Saharan Africa estimates that ∼26% of households affected by non-communicable diseases experience CHE when defined as 10% of total household expenditure—substantially higher than the ∼16.5% incidence observed across all health conditions combined (Eze *et al*., 2022). Meanwhile, a hospital-based study from Ethiopia found that 77.7% of cancer inpatients incurred CHE when measured as 40% of non-food expenditure, with CHE strongly associated with lower income and lack of insurance (Matebie *et al*., 2024). These comparisons suggest that our model’s implicit assumptions are conservative: real-world CHE from CC in Mozambique is likely higher, especially among the poorest, emphasizing the critical need for comprehensive treatment subsidies and support for indirect costs to protect vulnerable households.

Several limitations of this study need to be acknowledged. These relate to simplifying assumptions made around input parameters as well as the model structure. Firstly, national epidemiological information on age-specific CC incidence was available, but incidence rates were only based on the registry of two hospitals located in Beira in central Mozambique and Maputo in the south of Mozambique. The lack of data from other parts of the country and poor quality data on CC is likely to result in an underestimated number of CC cases, as observed in neighbouring Eswatini (Ginindza and Sartorius, 2018). Thus, the estimated impact of the vaccination programme is likely to be higher.

Secondly, data from a cohort study conducted in Dar es Salaam, Tanzania, informed the assumption that CC incidence is higher in the poorest quintiles. However, the reliability of this data is uncertain, as it was collected from a single hospital in Tanzania and has not been published in peer-reviewed literature. To account for potential variations, we explored different distributions of CC incidence across wealth quintiles. Our analysis showed that when CC risk is assumed to be uniform across all income groups and vaccination coverage follows a gradient favoring wealthier quintiles, the greatest health benefits accrue to the highest income group. This is because higher coverage in wealthier populations results in a larger absolute reduction in CC cases, even when baseline risk is the same across groups. However, this scenario - where the highest income quintile benefits the most - only occurs when coverage is skewed toward wealthier groups while all other factors remain constant. In reality, CC incidence is more likely to decrease with increasing income, a pattern observed in multiple countries (Chang *et al*., 2018). This suggests that, without targeted efforts to increase vaccination rates among lower-income groups, disparities in CC burden may persist or even widen.

Thirdly, the vaccination coverage data used in the status quo scenario, while based on reported figures, are subject to considerable uncertainty. Interviews revealed inaccuracies in the 2011 census data, the denominator for coverage calculations. Given that these coverage estimates were likely not adjusted prior to being shared, the actual coverage achieved in the program’s first year is likely lower than reported. While a more recent Post Enumeration Survey (PES) indicated a 3.7% undercount in the census, this does not fully address the potential inaccuracies in the older data used for the initial coverage calculations. Furthermore, coverage of the final dose reached 42.1% in 2021 (Kouassi *et al*., 2024), suggesting potential higher coverage rates in the future.

Therefore, to explore the sensitivity of our findings to coverage levels, we also analyzed scenarios with 50% and 90% coverage. The results across this range of coverage rates consistently demonstrate the value of investing in scaling up the program. This suggests that even with the inherent uncertainty surrounding the precise coverage achieved, the program’s impact and benefits to the healthcare system are substantial. While our model demonstrates the cost-effectiveness of the intervention across a range of scenarios, it is important to acknowledge that real-world implementation may involve scale-dependent cost variations. Our estimates assume vaccine distribution through fixed facilities, which may underestimate the additional costs required to reach remote or underserved populations. Outreach programs, for example, introduce extra logistical expenses, increasing the cost per fully immunized girl compared to fixed-facility delivery. Additionally, vaccination costs can vary depending on the chosen delivery strategy and the scale of implementation. Economies of scale could reduce per-dose costs in large-scale rollouts, whereas logistical challenges at higher coverage levels may drive costs up. Direct non-medical expenses, such as transportation, may also vary by wealth quintile and place of residence, influencing financial burden and access to care. Thus, the cost estimates presented in this study are conservative, as they do not fully account for these scale-dependent variations. Despite this, the consistent positive impact of increased coverage across plausible cost scenarios strengthens the case for expanding the vaccination program.

Fourthly, we adopted treatment cost and labor requirement estimates from Tanzania, which has a similar Healthcare Access and Quality Index (Fullman *et al*., 2018) and was therefore presumed to have comparable treatment capacities. This approach was necessary due to the lack of treatment cost data specific to Mozambique. However, the available data did not include uncertainty ranges, so all cost estimates were adjusted with a ±20% interval to reflect the high degree of uncertainty in this input value.

This model does not include the effects of HPV vaccination in reducing onward transmission. As such, the health impact of vaccination was under-estimated. Indeed there is strong evidence for vaccine cross-protection and herd effects from the HPV vaccine if coverage is 50% or above (Drolet *et al*., 2019). We also did not include the potential health benefits and costs associated with other preventive interventions, such as screening or treatment for precancerous lesions. Evaluations of combined strategies conducted in other countries concluded that, together, the interventions have the potential to eliminate CC (Brisson *et al*., 2020). Indirect costs, i.e., wages lost due to the inability of the patient or accompanying family members to work, were not estimated. These additional cost savings would further decrease CHE incidence. Further, household consumption only accounts for goods and services, i.e., the net of expenditure. Thus, CHE incidence estimates are possibly underestimated.

Due to limited data on medical resource requirements for CC patients and overall healthcare demand in Mozambique, we estimated only the labor saved as an example of the vaccine’s value to the health sector. In theory, reducing CC-related healthcare demands could allow resources to be redirected to other treatment services. However, in practice, healthcare resources are often over-committed, meaning that spare capacity may not necessarily become available. Patients who survive due to vaccination will continue to engage with healthcare services for other needs, which could offset some of the estimated resource savings. Future research could explore these dynamics further, including estimating additional resources saved, such as hospital bed days.

The ECEA framework used in this study to assess the distributional consequences of Mozambique’s HPV vaccination programme has previously been applied to evaluate the equity impact of other vaccines, including the HPV vaccine in other countries. This study adds to the existing literature by providing evidence on the health and economic impact of HPV vaccination specifically in Mozambique. The findings align with prior research, confirming that HPV infection disproportionately affects marginalized and lower-income populations due to limited access to screening and early detection (Driessen *et al*., 2015; Levin *et al*., 2015; Loganathan *et al*., 2016; Portnoy *et al*., 2021). Consequently, the greatest health benefits of the vaccination programme are concentrated in the lowest wealth quintiles. However, the financial benefits, in absolute terms, accrue more to wealthier households, given their greater ability to afford care and avoid catastrophic health expenditures. In relative terms, the financial relief may still be significant for lower-income households, given their higher risk of financial hardship from out-of-pocket medical expenses. While this analysis provides valuable insights into the distributional consequences of HPV vaccination, it does not quantify the programme’s impact on overall inequality in lifetime health. A distributional cost-effectiveness analysis (Dawkins *et al*., 2018) could complement this research by further exploring broader dimensions of the vaccine’s value and its impact on health disparities.

A key aspect of this modeling effort was collaboration with in-country partners to assess the availability and quality of local data. While data limitations posed significant challenges, the process itself provided valuable insights into why specific data are necessary for robust health policy modeling. In this way, this work highlighted critical data gaps and informing future data collection efforts. Additionally, engagement with Mozambican stakeholders played a role in shaping the research outcomes, including necessary model adaptations. This underscores the value of participatory modeling approaches in ensuring that analyses remain relevant and actionable within local contexts.

## 8 Conclusion

This study presents a comprehensive assessment of the HPV vaccination programme in Mozambique, considering its impact from a population health, health system, and patient perspective. The analysis of distributional effects across residence types and income groups is particularly relevant to Mozambique’s National Cancer Control Plan, which is currently under development (World Health Organization, 2022). The findings contribute to ongoing policy dialogues by providing evidence to support fair and efficient decision-making in HPV vaccination planning at both national and global levels.

## Author contributor statement

● Conception or design of the work: PC, AP
● Data collection: PC, AL, MM
● Data analysis and interpretation: PC, AL, MM
● Drafting the article: PC
● Critical revision of the article: PC, MM, TH, LC
● Final approval of the version to be submitted: PC, AP, AL, MM, NT, TH, LC

## Reflexivity statement

In conducting this modeling study, we acknowledge that our backgrounds, experiences, and perspectives have inevitably influenced the research process. The authors include five females and two males and span multiple levels of seniority. The team comprises two senior researchers specializing in health economics with a focus on vaccine impact, four social scientists with extensive experience in qualitative research, and three mathematical modellers of infectious diseases. Three of the authors are based in Mozambique at an institution actively engaged in policy dialogues on infectious disease interventions, including the HPV vaccination program. As mathematical modelers, health economists, and health policy and systems researchers, our disciplinary orientations and prior knowledge shaped how we interpreted data and conducted the overall analysis. Recognizing these influences, we sought to mitigate potential biases through collaborative engagement with local experts, iterative feedback, and reflexive consideration of how our perspectives informed the study’s direction and conclusions.

## Ethical Approval

Ethical approval for this type of study is not required by our institute.

## Data Availability

All data produced in the present study are available upon reasonable request to the authors

## Acknowledgements

An earlier version of this paper was presented at the International Papillomavirus Conference and a seminar at Imperial College London and the University of Nairobi, where we received valuable comments from participants. We also appreciate the input of study participants of preceding qualitative research, who shaped the research questions posed in this study and gave access to vaccine coverage data.

## Figure and Table Legends

Supplement 1 provides additional methodological details supporting the analysis presented in the manuscript, specifically on estimating household expenditures, cancer stage distribution, and access to cervical cancer (CC) treatment in Mozambique.

