## Supplement for "Health and economic impact of the human papillomavirus (HPV) vaccine in Mozambique"

### Supplement 1

**Outline of Goldie et al’s model**

A static cohort model previously developed by Goldie et al. (Goldie et al., 2008) was used. The model can be used to project the health impact of HPV vaccination at population level. The target cohort group(s) (e.g., 9-to-14-year-old girls) are tracked prospectively over their lifetime. In each time step, in this case year, the number of CC cases caused by HPV infection is simulated based on simplifying assumptions (duration and stage distribution of, and mortality from CC, HPV-16/18 serotype distribution in CC). The number of averted cases and deaths is estimated by subtracting the outcomes assuming a vaccine is introduced from the outcomes assuming no vaccine is introduced. This reduction is applied at population level, considering age-structured population prospects and background mortality. Population health outcomes (the number of averted CC cases by stage following the International Federation of Gynaecology and Obstetrics, or FIGO, system and averted CC deaths) are then aggregated over the target cohort(s)’ lifetime horizon.

**Assumptions made in the Extended Cost Effectiveness Analysis**

Annual household expenditure was derived by scaling monthly per capita consumption (Instituto Nacional de Estatística, 2021) (as a proxy for expenditure) multiplied by the average household size (Instituto Nacional de Estatística, 2021) to an annual average estimate. We applied a stage distribution of detected cancers to population subgroup-specific health outcomes to estimate the number of CC cases by stage. The stage distribution was informed by regional estimates of stage distribution at diagnosis that were derived by the Institute for Health Metrics and Evaluation and estimated for Mozambique by Canfell *et al* (Canfell et al., 2020).

The number of treatment-seeking CC cases by stage was assumed to vary by wealth quintile, depending on access to care. Access to care is influenced by: (1) available resources, i.e., the capacity to treat CC patients, (2) the number of people requiring CC treatment, which varies by wealth quintile, (3) and a distribution of having access to care across wealth quintiles. As of 2021, 900 CC patients could be treated per year at the Central Hospital Maputo (“Prevention, diagnosis and management of cancer in Mozambique,” 2021). We assume that a proportion of people go to the Central Private Hospital, which also has a radiology department and only patients in the highest wealth quintile seek healthcare here. Therefore, with an estimate of 5,325 CC patients in 2021 (Bray et al., 2018), the probability of having access to care can be estimated by wealth quintile. The proportion of patients seeking treatment at Central Private Hospital in Maputo (4.7%) (Gironés et al., 2020), is added to the probability of having access to care in the highest quintile. The distribution of access to treatment across wealth quintiles is informed by information on general care-seeking behaviour (Instituto Nacional de Estatística, 2021). We apply this distribution to the treatment capacity for CC patients in Mozambique. These estimates are likely to be a fair representation of the access to different types of healthcare in Mozambique as they are informed by a large Household Budget Survey conducted by the National Institute of Statistics of Mozambique with a sample size of 11,480 households (58,118 individuals). The authors of the technical report in which this estimate is published have noted there is high within- country variability in access to private healthcare, mostly concentrated in the southern region. Due to this provincial level heterogeneity and potential changes since the date of the survey (2014/15) Access to treatment assumptions are varied in sensitivity analysis. Treatment costs are assumed to be the same at both treatment facilities, the private and public hospitals.

Treatment costs were informed by a study conducted in Dar es Salaam, Tanzania (Nelson et al., 2016), which had a similar Healthcare Access and Quality Index score to Mozambique, (Fullman et al., 2018) supporting the assumption of comparable treatment capacities. Nelson et al. estimated average treatment costs of US$2963.60 per patient in 2022 (Nelson et al., 2016). We adopted these cost estimates as presented by Nelson et al. (Nelson et al., 2016), disaggregated by labor costs (for physicians, X-ray and ultrasound technologists, laboratory technologists, radiation technologists and team planning and initial consultation), supplies and equipment costs (X-ray and ultrasound, laboratory work, curative radiotherapy, chemotherapy, palliative radiotherapy), and inpatient accommodation costs. Costs were discounted at 3% annually.

Table 1 Average of cervical cancer incidence per 100,000 recorded in the Beira registry (2014-2017) and the Maputo registry (2015-2017) by age group.

| Agre Group | Cervical cancer incidence per 100,000 |
| --- | --- |
| 0-4 | 0 |
| 5-9 | 0 |
| 10-14 | 0.6 |
| 15-19 | 0 |
| 20-24 | 4 |
| 25-29 | 18.7 |
| 30-34 | 50.4 |
| 35-39 | 91.1 |
| 40-44 | 109.0 |
| 45-49 | 105.6 |
| 50-54 | 131.2 |
| 55-59 | 141.4 |
| 60-64 | 143.4 |
| 65-69 | 125.0 |
| 70-74 | 87.1 |
| 75-79 | 73.3 |
| 80-84 | 0 |
| 85-89 | 0 |
| 90-94 | 0 |
| 95-99 | 0 |

Fullman, N., Yearwood, J., Abay, S.M., Abbafati, C., Abd-Allah, F., Abdela, J., Abdelalim, A., Abebe, Z., Abebo, T.A., Aboyans, V., Abraha, H.N., Abreu, D.M.X., Abu-Raddad, L.J., Adane, A.A., Adedoyin, R.A., Adetokunboh, O., Adhikari, T.B., Afarideh, M., Afshin, A., Agarwal, G., Agius, D., Agrawal, A., Agrawal, S., Ahmad Kiadaliri, A., Aichour, M.T.E., Akibu, M., Akinyemi, R.O., Akinyemiju, T.F., Akseer, N., Al Lami, F.H., Alahdab, F., Al-Aly, Z., Alam, K., Alam, T., Alasfoor, D., Albittar, M.I., Alene, K.A., Al-Eyadhy, A., Ali, S.D., Alijanzadeh, M., Aljunid, S.M., Alkerwi, A., Alla, F., Allebeck, P., Allen, C., Alomari, M.A., Al-Raddadi, R., Alsharif, U., Altirkawi, K.A., Alvis-Guzman, N., Amare, A.T., Amenu, K., Ammar, W., Amoako, Y.A., Anber, N., Andrei, C.L., Androudi, S., Antonio, C.A.T., Araújo, V.E.M., Aremu, O., Ärnlöv, J., Artaman, A., Aryal, K.K., Asayesh, H., Asfaw, E.T., Asgedom, S.W., Asghar, R.J., Ashebir, M.M., Asseffa, N.A., Atey, T.M., Atre, S.R., Atteraya, M.S., Avila-Burgos, L., Avokpaho, E.F.G.A., Awasthi, A., Ayala Quintanilla, B.P., Ayalew, A.A., Ayele, H.T., Ayer, R., Ayuk, T.B., Azzopardi, P., Azzopardi-Muscat, N., Babalola, T.K., Badali, H., Badawi, A., Banach, M., Banerjee, A., Banstola, A., Barber, R.M., Barboza, M.A., Barker-Collo, S.L., Bärnighausen, T., Barquera, S., Barrero, L.H., Bassat, Q., Basu, S., Baune, B.T., Bazargan-Hejazi, S., Bedi, N., Beghi, E., Behzadifar, Masoud, Behzadifar, Meysam, Bekele, B.B., Belachew, A.B., Belay, S.A., Belay, Y.A., Bell, M.L., Bello, A.K., Bennett, D.A., Bennett, J.R., Bensenor, I.M., Berhe, D.F., Bernabé, E., Bernstein, R.S., Beuran, M., Bhalla, A., Bhatt, P., Bhaumik, S., Bhutta, Z.A., Biadgo, B., Bijani, A., Bikbov, B., Birungi, C., Biryukov, S., Bizuneh, H., Bolliger, I.W., Bolt, K., Bou-Orm, I.R., Bozorgmehr, K., Brady, O.J., Brazinova, A., Breitborde, N.J.K., Brenner, H., Britton, G., Brugha, T.S., Butt, Z.A., Cahuana-Hurtado, L., Campos-Nonato, I.R., Campuzano, J.C., Car, J., Car, M., Cárdenas, R., Carrero, J.J., Carvalho, F., Castañeda-Orjuela, C.A., Castillo Rivas, J., Catalá-López, F., Cercy, K., Chalek, J., Chang, H.-Y., Chang, J.-C., Chattopadhyay, A., Chaturvedi, P., Chiang, P.P.-C., Chisumpa, V.H., Choi, J.-Y.J., Christensen, H., Christopher, D.J., Chung, S.-C., Ciobanu, L.G., Cirillo, M., Colombara, D., Conti, S., Cooper, C., Cornaby, L., Cortesi, P.A., Cortinovis, M., Costa Pereira, A., Cousin, E., Criqui, M.H., Cromwell, E.A., Crowe, C.S., Crump, J.A., Daba, A.K., Dachew, B.A., Dadi, A.F., Dandona, L., Dandona, R., Dargan, P.I., Daryani, A., Daryani, M., Das, J., Das, S.K., Das Neves, J., Davis Weaver, N., Davletov, K., De Courten, B., De Leo, D., De Neve, J.-W., Dellavalle, R.P., Demoz, G., Deribe, K., Des Jarlais, D.C., Dey, S., Dharmaratne, S.D., Dhimal, M., Djalalinia, S., Doku, D.T., Dolan, K., Dorsey, E.R., Dos Santos, K.P.B., Doyle, K.E., Driscoll, T.R., Dubey, M., Dubljanin, E., Duncan, B.B., Echko, M., Edessa, D., Edvardsson, D., Ehrlich, J.R., Eldrenkamp, E., El-Khatib, Z.Z., Endres, M., Endries, A.Y., Eshrati, B., Eskandarieh, S., Esteghamati, A., Fakhar, M., Farag, T., Faramarzi, M., Faraon, E.J.A., Faro, A., Farzadfar, F., Fatusi, A., Fazeli, M.S., Feigin, V.L., Feigl, A.B., Fentahun, N., Fereshtehnejad, S.-M., Fernandes, E., Fernandes, J.C., Fijabi, D.O., Filip, I., Fischer, F., Fitzmaurice, C., Flaxman, A.D., Flor, L.S., Foigt, N., Foreman, K.J., Frostad, J.J., Fürst, T., Futran, N.D., Gakidou, E., Gallus, S., Gambashidze, K., Gamkrelidze, A., Ganji, M., Gebre, A.K., Gebrehiwot, T.T., Gebremedhin, A.T., Gelaw, Y.A., Geleijnse, J.M., Geremew, D., Gething, P.W., Ghadimi, R., Ghasemi Falavarjani, K., Ghasemi-Kasman, M., Gill, P.S., Giref, A.Z., Giroud, M., Gishu, M.D., Giussani, G., Godwin, W.W., Goli, S., Gomez-Dantes, H., Gona, P.N., Goodridge, A., Gopalani, S.V., Goryakin, Y., Goulart, A.C., Grada, A., Griswold, M., Grosso, G., Gugnani, H.C., Guo, Y., Gupta, Rahul, Gupta, Rajeev, Gupta, Tanush, Gupta, Tarun, Gupta, V., Haagsma, J.A., Hachinski, V., Hafezi-Nejad, N., Hailu, G.B., Hamadeh, R.R., Hamidi, S., Hankey, G.J., Harb, H.L., Harewood, H.C., Harikrishnan, S., Haro, J.M., Hassen, H.Y., Havmoeller, R., Hawley, C., Hay, S.I., He, J., Hearps, S.J.C., Hegazy, M.I., Heibati, B., Heidari, M., Hendrie, D., Henry, N.J., Herrera Ballesteros, V.H., Herteliu, C., Hibstu, D.T., Hiluf, M.K., Hoek, H.W., Homaie Rad, E., Horita, N., Hosgood, H.D., Hosseini, M., Hosseini, S.R., Hostiuc, M., Hostiuc, S., Hoy, D.G., Hsairi, M., Htet, A.S., Hu, G., Huang, J.J., Iburg, K.M., Idris, F., Igumbor, E.U., Ikeda, C., Ileanu, B.V., Ilesanmi, O.S., Innos, K., Irvani, S.S.N., Irvine, C.M.S., Islami, F., Jacobs, T.A., Jacobsen, K.H., Jahanmehr, N., Jain, R., Jain, S.K., Jakovljevic, M.B., Jalu, M.T., Jamal, A.A., Javanbakht, M., Jayatilleke, A.U., Jeemon, P., Jha, R.P., Jha, V., Jóúwiak, J., John, O., Johnson, S.C., Jonas, J.B., Joshua, V., Jürisson, M., Kabir, Z., Kadel, R., Kahsay, A., Kalani, R., Kar, C., Karanikolos, M., Karch, A., Karema, C.K., Karimi, S.M., Kasaeian, A., Kassa, D.H., Kassa, G.M., Kassa, T.D., Kassebaum, N.J., Katikireddi, S.V., Kaul, A., Kawakami, N., Kazanjan, K., Kebede, S., Keiyoro, P.N., Kemp, G.R., Kengne, A.P., Kereselidze, M., Ketema, E.B., Khader, Y.S., Khafaie, M.A., Khajavi, A., Khalil, I.A., Khan, E.A., Khan, G., Khan, M.N., Khan, M.A., Khanal, M.N., Khang, Y.-H., Khater, M.M., Khoja, A.T.A., Khosravi, A., Khubchandani, J., Kibret, G.D., Kiirithio, D.N., Kim, D., Kim, Y.J., Kimokoti, R.W., Kinfu, Y., Kinra, S., Kisa, A., Kissoon, N., Kochhar, S., Kokubo, Y., Kopec, J.A., Kosen, S., Koul, P.A., Koyanagi, A., Kravchenko, M., Krishan, K., Krohn, K.J., Kuate Defo, B., Kumar, G.A., Kumar, P., Kutz, M., Kuzin, I., Kyu, H.H., Lad, D.P., Lafranconi, A., Lal, D.K., Lalloo, R., Lam, H., Lan, Q., Lang, J.J., Lansingh, V.C., Lansky, S., Larsson, A., Latifi, A., Lazarus, J.V., Leasher, J.L., Lee, P.H., Legesse, Y., Leigh, J., Leshargie, C.T., Leta, S., Leung, J., Leung, R., Levi, M., Li, Y., Liang, J., Liben, M.L., Lim, L.-L., Lim, S.S., Lind, M., Linn, S., Listl, S., Liu, P., Liu, S., Lodha, R., Lopez, A.D., Lorch, S.A., Lorkowski, S., Lotufo, P.A., Lucas, T.C.D., Lunevicius, R., Lurton, G., Lyons, R.A., Maalouf, F., Macarayan, E.R.K., Mackay, M.T., Maddison, E.R., Madotto, F., Magdy Abd El Razek, H., Magdy Abd El Razek, M., Majdan, M., Majdzadeh, R., Majeed, A., Malekzadeh, R., Malhotra, R., Malta, D.C., Mamun, A.A., Manhertz, T., Manguerra, H., Mansournia, M.A., Mantovani, L.G., Manyazewal, T., Mapoma, C.C., Margono, C., Martinez-Raga, J., Martins, S.C.O., Martins-Melo, F.R., Martopullo, I., März, W., Massenburg, B.B., Mathur, M.R., Maulik, P.K., Mazidi, M., McAlinden, C., McGrath, J.J., McKee, M., Mehata, S., Mehrotra, R., Mehta, K.M., Mehta, V., Meier, T., Mejia-Rodriguez, F., Meles, K.G., Melku, M., Memiah, P., Memish, Z.A., Mendoza, W., Mengiste, D.A., Mengistu, D.T., Menota, B.G., Mensah, G.A., Meretoja, A., Meretoja, T.J., Mezgebe, H.B., Miazgowski, T., Micha, R., Milam, R., Millear, A., Miller, T.R., Mini, G., Minnig, S., Mirica, A., Mirrakhimov, E.M., Misganaw, A., Mitchell, P.B., Mlashu, F.W., Moazen, B., Mohammad, K.A., Mohammadibakhsh, R., Mohammed, E., Mohammed, M.A., Mohammed, S., Mokdad, A.H., Mola, G.L., Molokhia, M., Momeniha, F., Monasta, L., Montañez Hernandez, J.C., Moosazadeh, M., Moradi-Lakeh, M., Moraga, P., Morawska, L., Moreno Velasquez, I., Mori, R., Morrison, S.D., Moses, M., Mousavi, S.M., Mueller, U.O., Murhekar, M., Murthy, G.V.S., Murthy, S., Musa, J., Musa, K.I., Mustafa, G., Muthupandian, S., Nagata, C., Nagel, G., Naghavi, M., Naheed, A., Naik, G.A., Naik, N., Najafi, F., Naldi, L., Nangia, V., Nansseu, J.R.N., Narayan, K.V., Nascimento, B.R., Negoi, I., Negoi, R.I., Newton, C.R., Ngunjiri, J.W., Nguyen, G., Nguyen, L., Nguyen, T.H., Nichols, E., Ningrum, D.N.A., Nolte, E., Nong, V.M., Norheim, O.F., Norrving, B., Noubiap, J.J.N., Nyandwi, A., Obermeyer, C.M., Ofori-Asenso, R., Ogbo, F.A., Oh, I.-H., Oladimeji, O., Olagunju, A.T., Olagunju, T.O., Olivares, P.R., Oliveira, P.P.V.D., Olsen, H.E., Olusanya, B.O., Olusanya, J.O., Ong, K., Opio, J.N., Oren, E., Ortega-Altamirano, D.V., Ortiz, A., Ozdemir, R., Pa, M., Pain, A.W., Palone, M.R.T., Pana, A., Panda-Jonas, S., Pandian, J.D., Park, E.-K., Parsian, H., Patel, T., Pati, S., Patil, S.T., Patle, A., Patton, G.C., Paturi, V.R., Paudel, D., Pedroso, M.D.M., Pedroza, S.P., Pereira, D.M., Perico, N., Peterson, H., Petzold, M., Peykari, N., Phillips, M.R., Piel, F.B., Pigott, D.M., Pillay, J.D., Piradov, M.A., Polinder, S., Pond, C.D., Postma, M.J., Pourmalek, F., Prakash, S., Prakash, V., Prasad, N., Prasad, N.M., Purcell, C., Qorbani, M., Quintana, H.K., Radfar, A., Rafay, A., Rafiei, A., Rahimi, K., Rahimi-Movaghar, A., Rahimi-Movaghar, V., Rahman, M., Rahman, M.A., Rahman, S.U., Rai, R.K., Raju, S.B., Ram, U., Rana, S.M., Rankin, Z., Rasella, D., Rawaf, D.L., Rawaf, S., Ray, S.E., Razo-García, C.A., Reddy, P., Reiner, R.C., Reis, C., Reitsma, M.B., Remuzzi, G., Renzaho, A.M.N., Resnikoff, S., Rezaei, S., Rezai, M.S., Ribeiro, A.L., Rios Blancas, M.J., Rivera, J.A., Roever, L., Ronfani, L., Roshandel, G., Rostami, A., Roth, G.A., Rothenbacher, D., Roy, A., Roy, N., Ruhago, G.M., Sabde, Y.D., Sachdev, P.S., Sadat, N., Safdarian, M., Safiri, S., Sagar, R., Sahebkar, A., Sahraian, M.A., Sajadi, H.S., Salama, J., Salamati, P., Saldanha, R.D.F., Salimzadeh, H., Salomon, J.A., Samy, A.M., Sanabria, J.R., Sancheti, P.K., Sanchez-Niño, M.D., Santomauro, D., Santos, I.S., Santric Milicevic, M.M., Sarker, A.R., Sarrafzadegan, N., Sartorius, B., Satpathy, M., Savic, M., Sawhney, M., Saxena, S., Saylan, M.I., Schaeffner, E., Schmidhuber, J., Schmidt, M.I., Schneider, I.J.C., Schumacher, A.E., Schutte, A.E., Schwebel, D.C., Schwendicke, F., Sekerija, M., Sepanlou, S.G., Servan-Mori, E.E., Shafieesabet, A., Shaikh, M.A., Shakh-Nazarova, M., Shams-Beyranvand, M., Sharafi, H., Sharif-Alhoseini, M., Shariful Islam, S.M., Sharma, M., Sharma, R., She, J., Sheikh, A., Shfare, M.T., Shi, P., Shields, C., Shigematsu, M., Shinohara, Y., Shiri, R., Shirkoohi, R., Shiue, I., Shrime, M.G., Shukla, S.R., Siabani, S., Sigfusdottir, I.D., Silberberg, D.H., Silva, D.A.S., Silva, J.P., Silveira, D.G.A., Singh, J.A., Singh, L., Singh, N.P., Singh, V., Sinha, D.N., Sinke, A.H., Sisay, M., Skirbekk, V., Sliwa, K., Smith, A., Soares Filho, A.M., Sobaih, B.H.A., Somai, M., Soneji, S., Soofi, M., Sorensen, R.J.D., Soriano, J.B., Soyiri, I.N., Sposato, L.A., Sreeramareddy, C.T., Srinivasan, V., Stanaway, J.D., Stathopoulou, V., Steel, N., Stein, D.J., Stokes, M.A., Sturua, L., Sufiyan, M.B., Suliankatchi, R.A., Sunguya, B.F., Sur, P.J., Sykes, B.L., Sylaja, P., Tabarés-Seisdedos, R., Tadakamadla, S.K., Tadesse, A.H., Taffere, G.R., Tandon, N., Tariku, A.T., Taveira, N., Tehrani-Banihashemi, A., Temam Shifa, G., Temsah, M.-H., Terkawi, A.S., Tesema, A.G., Tesfaye, D.J., Tessema, B., Thakur, J., Thomas, N., Thompson, M.J., Tillmann, T., To, Q.G., Tobe-Gai, R., Tonelli, M., Topor-Madry, R., Topouzis, F., Torre, A., Tortajada, M., Tran, B.X., Tran, K.B., Tripathi, A., Tripathy, S.P., Troeger, C., Truelsen, T., Tsoi, D., Tudor Car, L., Tuem, K.B., Tyrovolas, S., Uchendu, U.S., Ukwaja, K.N., Ullah, I., Updike, R., Uthman, O.A., Uzochukwu, B.S.C., Valdez, P.R., Van Boven, J.F.M., Varughese, S., Vasankari, T., Violante, F.S., Vladimirov, S.K., Vlassov, V.V., Vollset, S.E., Vos, T., Wagnew, F., Waheed, Y., Wallin, M.T., Walson, J.L., Wang, Y., Wang, Y.-P., Wassie, M.M., Weaver, M.R., Weiderpass, E., Weintraub, R.G., Weiss, J., Weldegwergs, K.G., Werdecker, A., West, T.E., Westerman, R., White, R.G., Whiteford, H.A., Widecka, J., Winkler, A.S., Wiysonge, C.S., Wolfe, C.D., Wondimkun, Y.A., Workicho, A., Wyper, G.M.A., Xavier, D., Xu, G., Yan, L.L., Yano, Y., Yaseri, M., Yimer, N.B., Yin, P., Yip, P., Yirsaw, B.D., Yonemoto, N., Yonga, G., Yoon, S.-J., Yotebieng, M., Younis, M.Z., Yu, C., Zadnik, V., Zaidi, Z., Zaki, M.E.S., Zaman, S.B., Zamani, M., Zenebe, Z.M., Zhou, M., Zhu, J., Zimsen, S.R.M., Zipkin, B., Zodpey, S., Zuhlke, L.J., Murray, C.J.L., Lozano, R., 2018. Measuring performance on the Healthcare Access and Quality Index for 195 countries and territories and selected subnational locations: a systematic analysis from the Global Burden of Disease Study 2016. The Lancet 391, 2236–2271. https://doi.org/10.1016/S0140-6736(18)30994-2

Gironés, A.L., Belvis, F., Julià, M., Benach, J., 2020. Health care inequalities in Mozambique: needs, access, barriers and quality of care.

Instituto Nacional de Estatística, 2021. Inquérito sobre orcamento familiar - IOF 2019/20.

Nelson, S., Kim, J., Wilson, F.A., Soliman, A.S., Ngoma, T., Kahesa, C., Mwaiselage, J., 2016. Cost-Effectiveness of Screening and Treatment for Cervical Cancer in Tanzania: Implications for other Sub-Saharan African Countries. Value Health Reg. Issues 10, 1–6. https://doi.org/10.1016/j.vhri.2016.03.002

Prevention, diagnosis and management of cancer in Mozambique, 2021.
